# Doubling dolutegravir dosage reduces the viral reservoir in ART-treated people with HIV

**DOI:** 10.1101/2025.04.15.25325914

**Authors:** Céline Fombellida-Lopez, Aurelija Valaitienė, Lee Winchester, Nathalie Maes, Patricia Dellot, Céline Vanwinge, Aurélie Ladang, Etienne Cavalier, Fabrice Susin, Dolores Vaira, Marie-Pierre Hayette, Catherine Reenaers, Michel Moutschen, Courtney V. Fletcher, Alexander O. Pasternak, Gilles Darcis

## Abstract

Whether antiretroviral therapy (ART) is always completely suppressive, or HIV might continue to replicate at low levels despite ART in some people with HIV (PWH), is still debated. Here, we intensified the ART regimen by doubling dolutegravir (DTG) dosage and investigated the impact of this strategy on HIV blood and tissue reservoirs, immune activation, and inflammation. Twenty HIV-infected adults, who had received a triple ART consisting of 50mg DTG/600 mg abacavir/300 mg lamivudine pre-intensification and had been suppressed on ART for at least two years, were enrolled in a phase 2 randomized clinical trial. Half of them received an additional 50 mg of DTG for a period of 84 days. As expected, plasma and tissue DTG concentrations significantly increased during the study period in the intensified group but not in the control group. Accordingly, significant decreases in total HIV DNA, intact HIV DNA, and cell-associated unspliced (US) HIV RNA in PBMCs, as well as in the US RNA/total DNA ratio, were observed in the intensified group but not in the control group. Intensification also modestly reduced markers of immune activation and exhaustion but had no measurable impact on systemic or tissue inflammation. Together with this, intensification resulted in a temporary decrease in the CD4/CD8 ratio that returned to baseline by day 84. Our results strongly suggest that the pre-intensification ART regimen was not completely suppressive. If confirmed in larger clinical trials, these results could have an impact on the clinical management of PWH and HIV curative strategies.

## Introduction

Human immunodeficiency virus (HIV) remains a major global public health issue, with an estimated 39.9 million people with HIV (PWH) at the end of 2023. Out of them, 30.7 million (77%) were accessing antiretroviral therapy (ART) (https://www.unaids.org/en/resources/fact-sheet). ART suppresses plasma viral load to undetectable levels, restores immune function, and eliminates the risk of developing AIDS (1). Current ART usually consists of one or two nucleotide or nucleoside reverse transcriptase inhibitors and the second or third drug of another class (e.g., a non-nucleoside reverse transcriptase inhibitor, a protease inhibitor (PI), or an integrase strand transfer inhibitor (INSTI)) (https://eacs.sanfordguide.com/). The mechanism of action of ART is interfering with different steps of the viral replication cycle, such as reverse transcription, integration, or viral particle maturation. Importantly, ART only blocks the infection of new cells and does not inhibit HIV gene expression in cells that were infected prior to ART initiation or in the progeny of such cells (2).

ART is not curative, and the main barrier to an HIV cure is the presence of latent viral reservoirs that arise due to the ability of HIV to integrate its genome into the host chromosome and persist in a latent form in some of the cells it infects, such as memory CD4+ T cells (3, 4). These reservoir cells harbour stably integrated and replication-competent provirus that can reactivate and, after cessation of ART, fuel a rapid viral rebound (5). HIV reservoirs persist lifelong, slowly decaying over time (6–8). Several mechanisms have been described to explain the long-term persistence of HIV reservoirs, including the infection of long-lived CD4+ T cells and clonal expansion by homeostatic or antigen-driven cellular proliferation or by HIV integration in or near genes that promote cellular proliferation (9, 10). However, if therapy does not prevent all new infections, then replenishment by *de novo* virus replication provides an additional potential mechanism of reservoir persistence (11).

Whether ART is fully suppressive is a topic of considerable and long-standing debate (12). Evidence of residual replication in ART-suppressed PWH has been previously presented (13–16), but other groups published reports that contest this notion (17–20). One strong argument against ongoing viral replication is the absence of detectable virus evolution on ART. However, if residual replication occurs intermittently and at a low level, with short infection chains continuously arising and terminating, then nucleotide substitutions are expected to be sporadic and not linked by temporal structure, unless the sampling is very deep and intensive (13, 21). In principle, failure to detect virus evolution does not form an unequivocal proof that residual viral replication does not occur at all: absence of evidence is not evidence of absence. Indeed, Paryad-Zanjani et al. recently developed a mathematical model to track the viral dynamics in lymph nodes (LNs) after the initiation of ART (22), which suggested that the methods used by the previous studies to detect genetic divergence were not sufficiently sensitive and that these studies would have been unlikely to detect residual replication.

Over time, this persistent low-level residual replication may contribute to chronic immune activation and inflammation in ART-treated PWH (23). Constant stimulation of the immune system could lead to immune exhaustion and premature immune senescence, resulting, over the years, in the development of non-AIDS co-morbidities, such as cardiovascular disease (24). This replication may occur in anatomical sanctuaries/compartments, notably the LNs and the gut-associated lymphoid tissue (GALT), two sites massively infected by HIV (25). Lower penetration of antiretroviral drugs to these tissues reduces the local drug concentrations and, consequently, their effectiveness in preventing HIV replication (26–28).

One way to investigate the residual replication is to intensify a standard ART regimen and measure the impact of this intensification on latent reservoirs, inflammation, and immune activation. If a reduction of a virological parameter is observed, it would strongly suggest that the ART regimen prior to intensification did not completely prevent virus replication. Several groups have conducted ART intensification trials, either by adding a CCR5 antagonist (maraviroc) or an INSTI (raltegravir or dolutegravir (DTG)) to a standard three-drug regimen. Some of these trials have indeed revealed residual virus replication at baseline, as treatment intensification caused a decrease in immune activation, a decrease in cell-associated HIV RNA in the ileum, and a transient increase in 2-long terminal repeat (2-LTR) circles (29–38). However, other groups have failed to demonstrate any effects of ART intensification on virological parameters (39–45).

In summary, is still unclear whether ongoing viral replication can occur in the presence of ART. The consequences of residual replication for the HIV curative strategies are important since approaches aimed at eliminating viral reservoirs such as latency reversal (“shock-and-kill”) (46) are not expected to be fully effective if the virus continues to replicate despite treatment. In the worst-case scenario, the residual replication could lead to significant replenishment of the HIV reservoir upon latency reversal, compensating for any reservoir depletion achieved by this strategy. Therefore, understanding the phenomenon of residual replication and developing strategies to eliminate it are key elements in the quest to cure PWH.

Here, we intensified ART in PWH who had been suppressed on ART for at least two years, by doubling the dosage of DTG, an antiretroviral drug that had already been part of the regimen pre-intensification. We investigated the impact of this intensification on HIV blood and tissue latent reservoirs, immune activation, and inflammation.

## Results

### Study design and participants

Between February and July 2019, twenty ART-suppressed participants receiving a triple antiretroviral regimen consisting of 50 mg DTG, 600 mg abacavir (ABC), and 300 mg lamivudine (3TC) daily were enrolled into two groups as follows: 10 participants continued to receive this regimen (control group) and 10 participants received an additional 50 mg of DTG daily for 84 days (intensified group) as shown in Figure S1. Study participants were mostly Caucasian males and the median age was 52 years. The median cumulative time of viral suppression on ART was 7.4 years and participants had maintained continuous viral suppression for a median of 3.9 years before the study. The clinical characteristics of the participants at baseline are shown in Table 1.

**Table 1.** Clinical characteristics of participants at baseline.

|  | All<br>(n=20) | Control group<br>(n=10) | DTG group<br>(n=10) | p<sup>a</sup> |
| --- | --- | --- | --- | --- |
| Sex, male | 19 (95.0)<sup>b</sup> | 9 (90.0) | 10 (100.0) | 1.00 |
| Age, years | 52 (43-60; 25-73) | 52 (40-62; 25-73) | 51 (46-59; 36-66) | 0.94 |
| BMI, kg/m<sup>2</sup> | 24 (22-26; 18-36) | 25 (22-27; 19-30) | 23 (22-26; 18-36) | 0.62 |
| Ethnicity |  |  |  |  |
| Caucasian | 17 (85.0) | 9 (90.0) | 8 (80.0) | 1.00 |
| African | 2 (10.0) | 1 (10.0) | 1 (10.0) |  |
| Maghrebi | 1 (5.0) | 0 (0.0) | 1 (10.0) |  |
| HLA typing B57/01, negative (n=19) | 19 (100.0) | 9 (100.0) | 10 (100.0) | - |
| Smoking, smoker or ex-smoker | 10 (50.0) | 4 (40.0) | 6 (60.0) | 0.66 |
| Time since first positive HIV serology, years | 12.6 (6.8-18.2; 3.5-25.4) | 9.8 (7.3-13.3; 3.5-25.4) | 15.6 (7.6-18.5; 3.5-21.5) | 0.58 |
| Cumulative time of untreated HIV infection, years | 0.9 (0.1-3.5; 0.0-17.2) | 1.8 (0.1-5.0; 0.0-17.2) | 0.6 (0.1-2.1; 0.0-6.1) | 0.47 |
| Cumulative time of viral suppression, years | 7.4 (4.8-9.2; 2.9-16.5) | 5.6 (4.7-7.7; 3.3-16.2) | 7.9 (5.8-12.4; 2.9-16.5) | 0.32 |
| Continuous time of viral suppression before study, years | 3.9 (3.3-5.8; 0.4-10.2) | 4.4 (3.7-6.9; 3.3-10.2) | 3.4 (3.2-4.9; 0.4-7.9) | 0.12 |
| Time on DTG-containing ART regimen before study, years | 3.5 (3.4-3.7; 2.2-3.9) | 3.5 (3.4-3.8; 2.2-3.9) | 3.5 (3.3-3.5; 2.7-3.8) | 0.41 |
| Nadir CD4+ count, cells/mm<sup>3</sup> | 277 (147-410; 24-747) | 277 (205-340; 24-521) | 292 (136-508; 30-747) | 0.65 |
| First measured plasma viral load, log<sub>10</sub> HIV RNA copies/mL | 4.52 (3.91-5.35; 2.48-6.60) | 4.52 (4.36-4.96; 3.00-6.60) | 4.55 (3.69-5.38; 2.48-5.70) | 0.85 |
| CD4+ count, cells/mm<sup>3</sup> | 775 (659-1144; 225-1383) | 732 (672-970; 225-1267) | 886 (668-1217; 492-1383) | 0.39 |
| CD8+ count, cells/mm<sup>3</sup> | 745 (637-1288; 377-2017) | 723 (535-931; 377-2017) | 939 (690-1310; 501-1853) | 0.39 |
| CD4/CD8 ratio | 0.94 (0.66-1.47; 0.32-2.24) | 1.00 (0.64-1.36; 0.32-1.82) | 0.93 (0.69-1.56; 0.35-2.24) | 0.82 |
| Total HIV DNA in PBMCs, copies/10<sup>6</sup> cells | 175.5 (70.0-467.3; 15.1-1090) | 99.4 (37.4-231.5; 15.1-333.0) | 421.0 (139.5-764.3; 61.8-1090) | 0.015 |
| Intact HIV DNA in PBMCs, copies/10<sup>6</sup> cells | 55.2 (19.2-87.0; 5.17-624.2) | 29.5 (16.2-97.1; 10.3-624.2) | 58.6 (44.7-144.7; 5.17-292.6) | 0.36 |
| US HIV RNA in PBMCs, copies/μg total RNA | 92.1 (13.3-290.3; 2.53-1580) | 72.2 (12.2-253.8; 6.18-486) | 124.6 (17.0-591.0; 2.53-1580) | 0.58 |
| US RNA/total DNA ratio in PBMCs | 0.40 (0.07-1.47; 0.03-3.96) | 1.21 (0.11-2.81; 0.04-3.96) | 0.33 (0.06-0.70; 0.03-2.73) | 0.21 |
| Total HIV DNA in rectal tissue, copies/10<sup>6</sup> cells | 477.0 (271.3-971.5; 7.27-1720) | 547.0 (291.0-1215; 53.4-1720) | 403.0 (194.6-952.5; 7.27-1510) | 0.44 |
| Plasma DTG concentration, ng/mL | 3287 (2602-5087; 237-6593) | 3560 (2953-4455; 1969-5536) | 3266 (2245-5983; 237-6593) | 0.82 |
| Tissue DTG concentration, ng/g | 634 (533-830; 303-1810) | 737 (534-852; 482-1810) | 581 (535-714; 303-1037) | 0.44 |
| Plasma 3TC concentration, ng/mL | 316 (148-731; 50-1616) | 431 (214-941; 102-1616) | 246 (117-439; 50-1156) | 0.26 |
| Tissue 3TC concentration, ng/g | 2114 (1417-2345; 90-4495) | 2193 (1590-2784; 1215-4495) | 2114 (1233-2317; 900-3187) | 0.50 |
| CDC classification |  |  |  | - |
| A1 | 4 (20.0) | 1 (10.0) | 3 (30.0) |  |
| A2 | 9 (45.0) | 6 (60.0) | 3 (30.0) |  |
| A3 | 3 (15.0) | 1 (10.0) | 2 (30.0) |  |
| B3 | 1 (5.0) | 0 (0.0) | 1 (10.0) |  |
| C3 | 3 (15.0) | 2 (20.0) | 1 (10.0) |  |
| HBV status |  |  |  | - |
| Immune | 12 (60.0) | 8 (80.0) | 4 (40.0) |  |
| Non-immune, not infected | 5 (25.0) | 2 (20.0) | 3 (30.0) |  |
| Isolated HBc Ab | 2 (10.0) | 0 (0.0) | 2 (20.0) |  |
| Cured Hepatitis B | 1 (5.0) | 0 (0.0) | 1 (10.0) |  |
| HCV status |  |  |  | 1.00 |
| Not Infected | 19 (95.0) | 10 (10.0) | 9 (90.0) |  |
| Recovered | 1 (5.0) | 0 (0.0) | 1 (10.0) |  |

<sup>a</sup> Mann-Whitney tests were used for continuous variables and Fisher's exact tests were used for categorical variables.
<sup>b</sup> Data are medians (interquartile ranges, followed by ranges) for continuous variables and numbers (percentages) for discrete variables.

### ART intensification increases plasma and tissue DTG concentrations

To assess if the desired experimental conditions were achieved, we monitored the concentrations of DTG and 3TC, two antiretroviral drugs that were part of the ART regimen (Figure 1, Figure S2). Baseline plasma and rectal tissue DTG and 3TC concentrations were not significantly different between the intensified and control groups (Table 1). At median, baseline DTG concentration in the rectal tissue was 16.7% (interquartile range (IQR), 15.1%-24.6%) of the corresponding DTG plasma concentration, strikingly close to the value (17%) reported previously (47). As expected, plasma and tissue DTG concentrations significantly increased between days 0 and 84 in the intensified group but not in the control group (Figures 1A, 1B). In fact, DTG concentrations almost exactly doubled in plasma (median fold increase, 1.97-fold) and more than doubled in tissue (median fold increase, 2.48-fold) on day 84, as compared to day 0, in the intensified group. However, at day 84, tissue DTG concentration in the intensified group was still much lower (median, 20.0%; IQR, 17.3%-22.3%) than the corresponding DTG plasma concentration, reflecting limited penetration of DTG into the tissue. Plasma and tissue 3TC concentrations did not change on day 84 compared to day 0 in both groups, which was expected as the dosage of 3TC was not increased in any group (Figures 1C, 1D). However, we did observe a transient increase in the plasma DTG concentration in the control group and in the plasma 3TC concentration in both groups on days 1 and 56 of the study (Figure S2), with the concentrations returning to baseline levels by day 84. One possible explanation for this effect could be an increased adherence of the participants to their ART regimens during the study (“white coat compliance”) (48).

**Figure 1.**
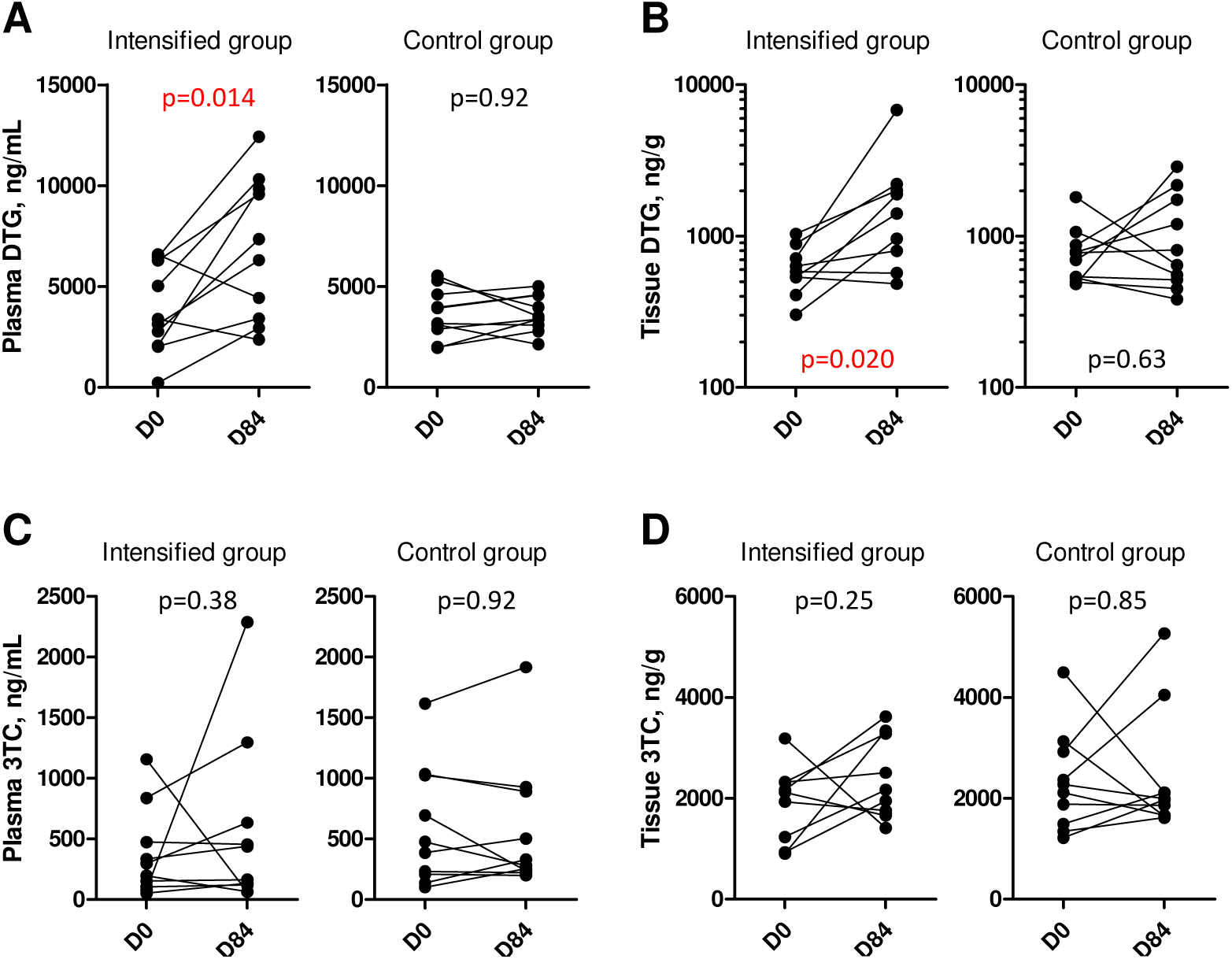
Antiretroviral drug concentrations in plasma and rectal tissue. Plasma (A) and tissue (B) concentrations of DTG, as well as plasma (C) and tissue (D) concentrations of 3TC, were compared between days 0 (D0) and 84 (D84) in intensified and control groups. Wilcoxon tests were used to calculate statistical significance.

### ART intensification reduces HIV reservoir

To investigate the effect of treatment intensification on the viral reservoir, we longitudinally measured total HIV DNA and cell-associated unspliced (US) HIV RNA in peripheral blood mononuclear cells (PBMCs) on days 0, 1, 28, 56, and 84 of the study. In addition, we measured intact HIV DNA in PBMCs and total HIV DNA in rectal biopsies on days 0 and 84 of the study. Strikingly, we observed significant longitudinal decreases in total HIV DNA, US RNA, and HIV transcription level per provirus (US RNA/total DNA ratio) in PBMCs in the intensified group (p=0.0022, p<0.001, and p<0.001, respectively) but not in the control group, with significant differences in the relative changes from baseline for two of these three markers between the groups (p<0.001, p=0.010, and p=0.090, respectively) (Figures 2A-2C, Figures S3-S5). While total HIV DNA in the intensified group transiently decreased in the first 28 days of the study (median (IQR) fold decrease from day 0 to day 28, 2.1 (1.1-4.7) fold, p=0.027) and increased afterwards (Figure 2A, Figure S3), US RNA and US RNA/total DNA ratio in the intensified group continued to decrease throughout the whole study period (Figures 2B-2C, Figures S4, S5). By day 84, US RNA and US RNA/total DNA ratio have decreased from day 0 by medians (IQRs) of 5.1 (3.3-6.4) and 4.6 (3.1-5.3) fold, respectively (p=0.016 for both markers). In comparison, in the control group these markers remained at the baseline level or even increased (Figures 2A-2C). These effects were confirmed by measurement of intact HIV DNA in PBMCs that was quantified by the intact proviral DNA assay (IPDA) (49). We measured a significant decrease in the intact HIV DNA between days 0 and 84 in the intensified group (median (IQR) fold decrease, 2.2 (1.3-3.1) fold, p=0.039) but not in the control group (Figure 2D). In contrast, total HIV DNA in rectal tissue did not change between days 0 and 84 in both groups (Figure 2E). Taken together, these results indicate that the intensification reduced the HIV reservoir in the peripheral blood of ART-treated PWH.

**Figure 2.**
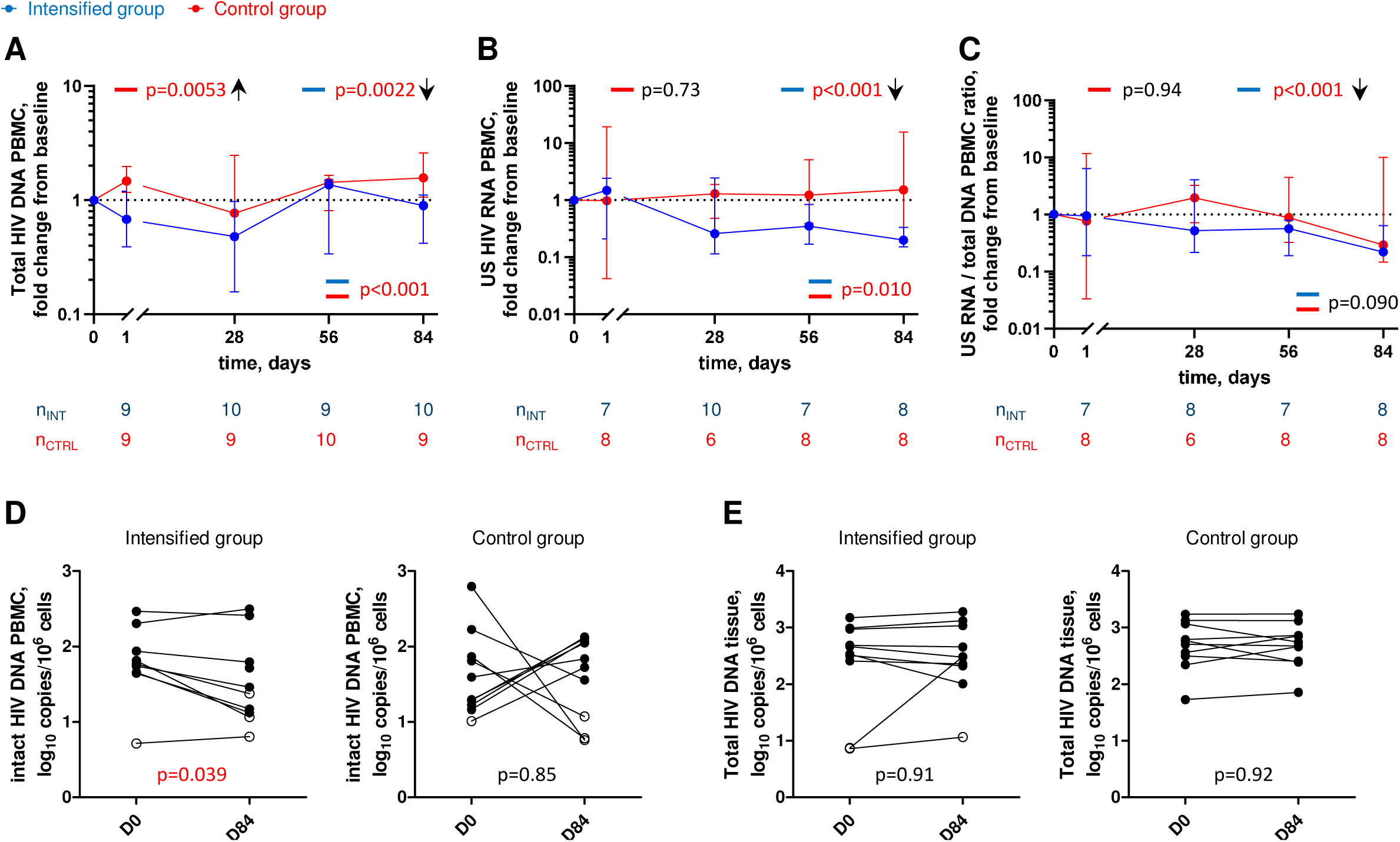
Longitudinal dynamics of HIV reservoir markers. (A-C) Fold change of total HIV DNA in PBMCs (A), US HIV RNA in PBMCs (B), and US RNA/total DNA ratios (C) from baseline on days 1, 28, 56, and 84 of the study in the intensified (blue) and control (red) groups. Median values and IQRs are shown. Linear mixed-effects modelling on log_10_-transformed values was used to calculate statistical significance. P values at the bottom of the graphs show the significance of between-group comparisons and those on top of the graphs show the significance of comparisons of the changes from baseline with zero in each group separately (intercept-only analysis). An upward or downward facing arrow next to a p value indicates a statistically significant increase or decrease from baseline, respectively. Participant numbers in both groups per time point are indicated below the graphs. (D, E) Comparisons of intact HIV DNA in PBMCs (D) and of total HIV DNA in rectal tissue (E) between days 0 (D0) and 84 (D84) in the intensified and control groups. Open circles depict undetectable values, assigned the values corresponding to 50% of the assay detection limits. Wilcoxon tests were used to calculate statistical significance. All p values are marked red if significant (<0.05).

### ART intensification modestly reduces markers of immune activation and exhaustion

To detect if intensification had an impact on immune activation and exhaustion, we performed longitudinal flow cytometry analysis to assess the expression of cell surface markers of activation (HLA-DR, CD38) and exhaustion (PD-1, TIGIT) on CD4+ and CD8+ T cells (Figure 3, Figure S6). A transient but significant increase in the percentage of CD4+ cells expressing PD-1 (p=0.022), and a significant decrease in the percentage of CD8+ cells expressing CD38 (p=0.034) were measured in the control group. A decrease in the percentage of CD4+ cells expressing TIGIT was measured in the intensified group (p=0.031), with a significant difference compared to the control group (p=0.048). We also measured a modest yet significant decrease in the percentage of CD8+ cells co-expressing CD38 and HLA-DR in the intensified group (p=0.049) (Figure 3). Overall, these results indicate that intensification modestly reduced markers of immune activation and exhaustion.

**Figure 3.**
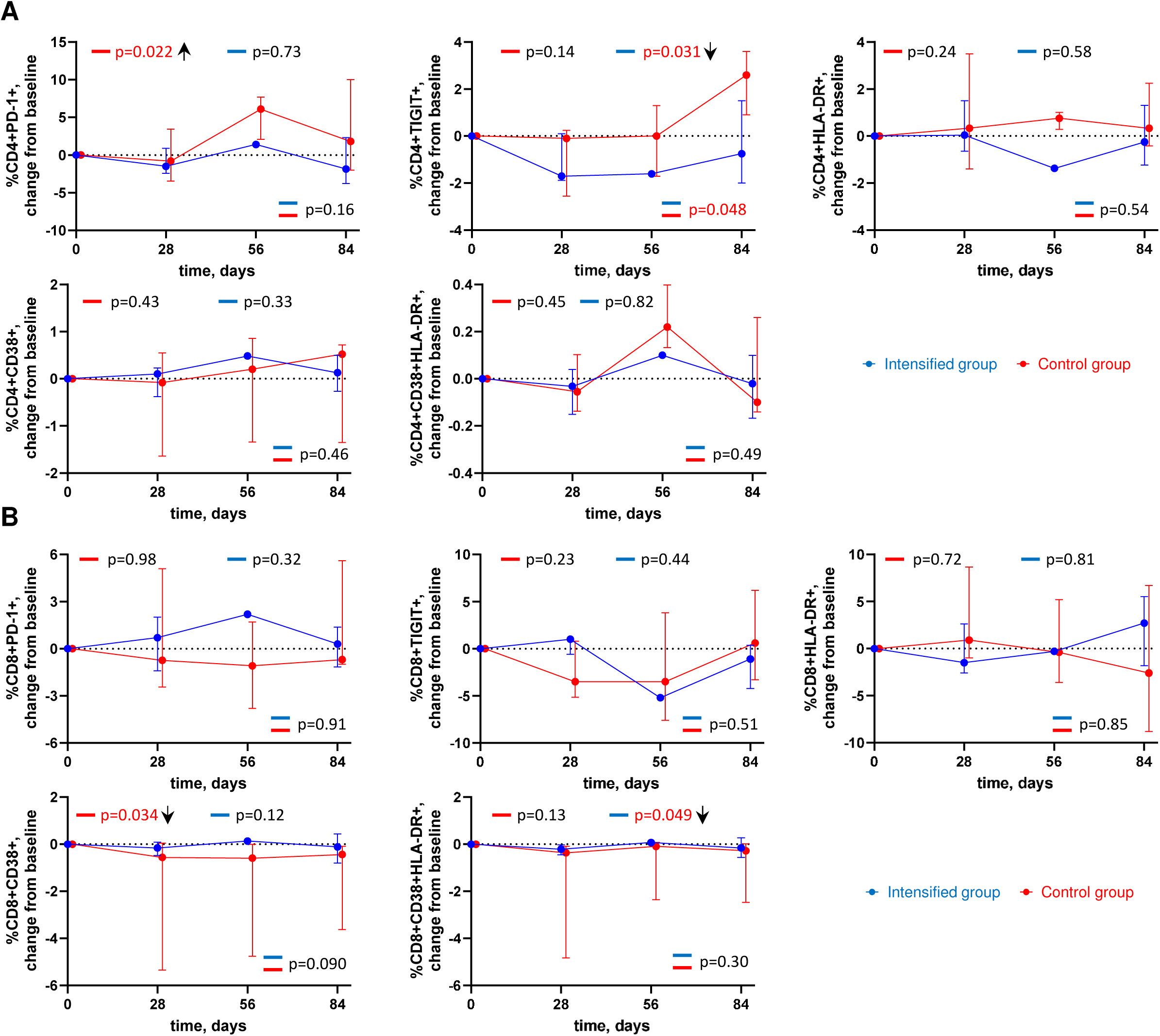
Longitudinal dynamics of cellular markers of immune activation and exhaustion. Graphs show changes of. (A) CD4+ cell markers and (B) CD8+ cell markers from baseline on days 1, 28, 56, and 84 of the study in the intensified (blue) and control (red) groups. Median values and IQRs are shown. Linear mixed-effects modelling was used to calculate statistical significance. P values at the bottom of the graphs show the significance of between-group comparisons and those on top of the graphs show the significance of comparisons of the changes from baseline with zero in each group separately (intercept-only analysis). An upward or downward facing arrow next to a p value indicates a statistically significant increase or decrease from baseline, respectively. P values are marked red if significant (<0.05).

### ART intensification does not impact systemic or tissue inflammation

To investigate the effect of intensification on systemic and tissue inflammation, we quantified several inflammatory cytokines in plasma and in rectal tissue. In plasma, we longitudinally measured the levels of IL-1β, IL-6, IFN-γ, IL-17 α, and TNF-α, and in rectal tissue, we measured expression of the same cytokines by quantifying their mRNA levels at days 0 and 84. We observed a significant increase in plasma IL-6 levels in the control group (p=0.049). Apart from that, no significant change from baseline for any cytokine was observed for both groups in both plasma and tissue (Figure 4, Figure S7). Overall, these results indicate that intensification did not measurably impact systemic or tissue inflammation.

**Figure 4.**
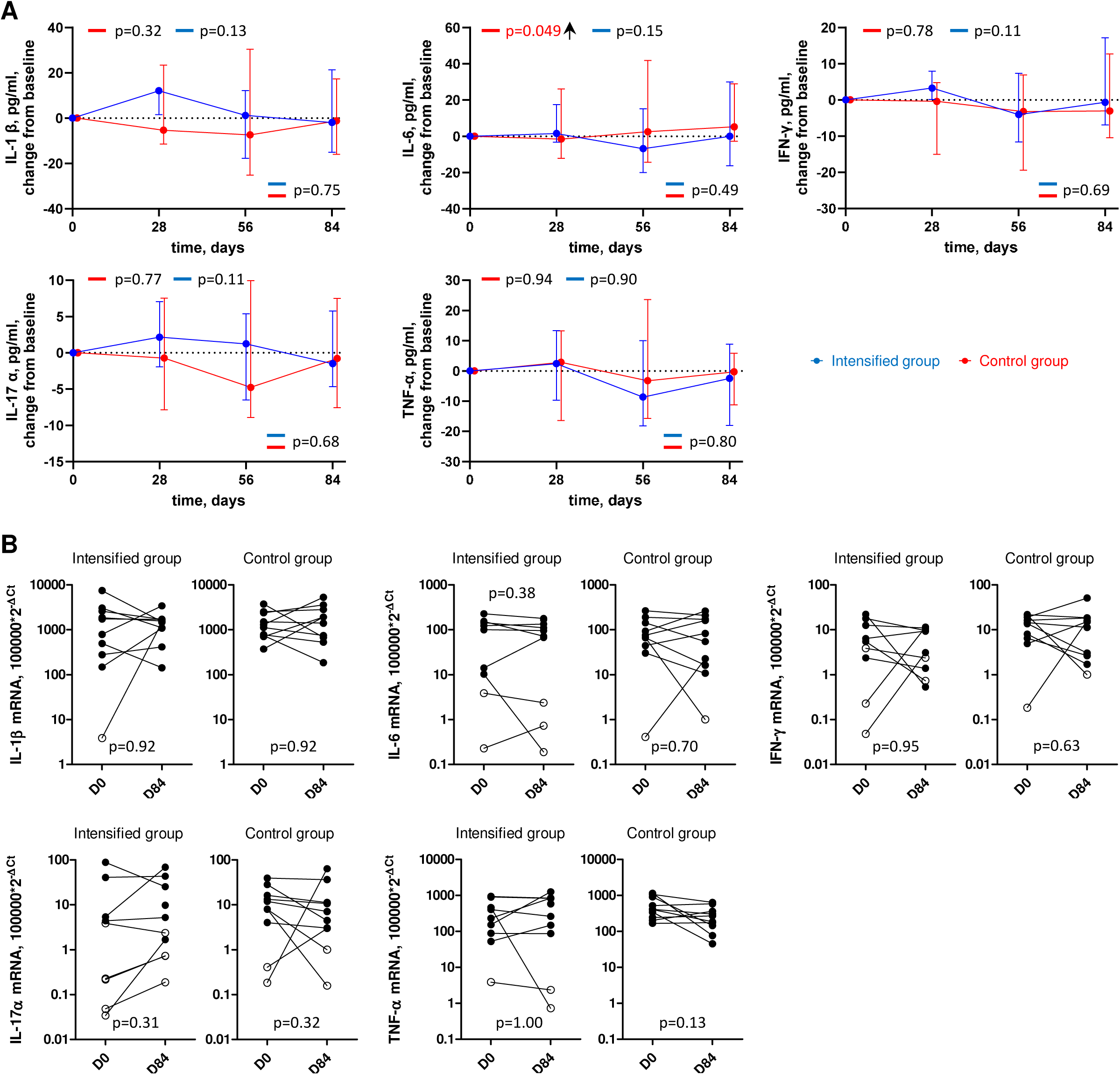
Longitudinal dynamics of inflammation markers in plasma and tissue. (A) Plasma inflammation markers. Graphs show changes of markers from baseline on days 1, 28, 56, and 84 of the study in the intensified (blue) and control (red) groups. Median values and IQRs are shown. Linear mixed-effects modelling was used to calculate statistical significance. P values at the bottom of the graphs show the significance of between-group comparisons and those on top of the graphs show the significance of comparisons of the changes from baseline with zero in each group separately (intercept-only analysis). An upward or downward facing arrow next to a p value indicates a statistically significant increase or decrease from baseline, respectively. P values are marked red if significant (<0.05). (B) Tissue inflammation markers. Markers were compared between days 0 (D0) and 84 (D84) in the intensified and control groups. Open circles depict undetectable values, assigned the values corresponding to 50% of the assay detection limits. Wilcoxon tests were used to calculate statistical significance.

### ART intensification leads to a transient decrease in the CD4/CD8 ratio

We longitudinally measured CD4 and CD8 T-cell counts, CD4/CD8 ratio, as well as plasma levels of soluble CD14 (sCD14), a marker of monocyte/macrophage activation, and of C-reactive protein (CRP), an inflammation marker (Figure 5, Figure S8). No significant changes were observed in the CD4 and CD8 counts in both groups, however we measured a transient but significant decrease in the CD4/CD8 ratio in the intensified group (p=0.0081) that returned to the baseline level by day 84 (Figure 5). No change in the CD4/CD8 ratio was observed in the control group. We also observed a significant decrease in the sCD14 level in the control group (p<0.001) and a trend toward an increase in the intensified group (p=0.079), which resulted in a significant difference between the groups (p<0.001). Finally, we observed a small but significant increase in the CRP level in the control group (p=0.045) but not in the intensified group.

**Figure 5.**
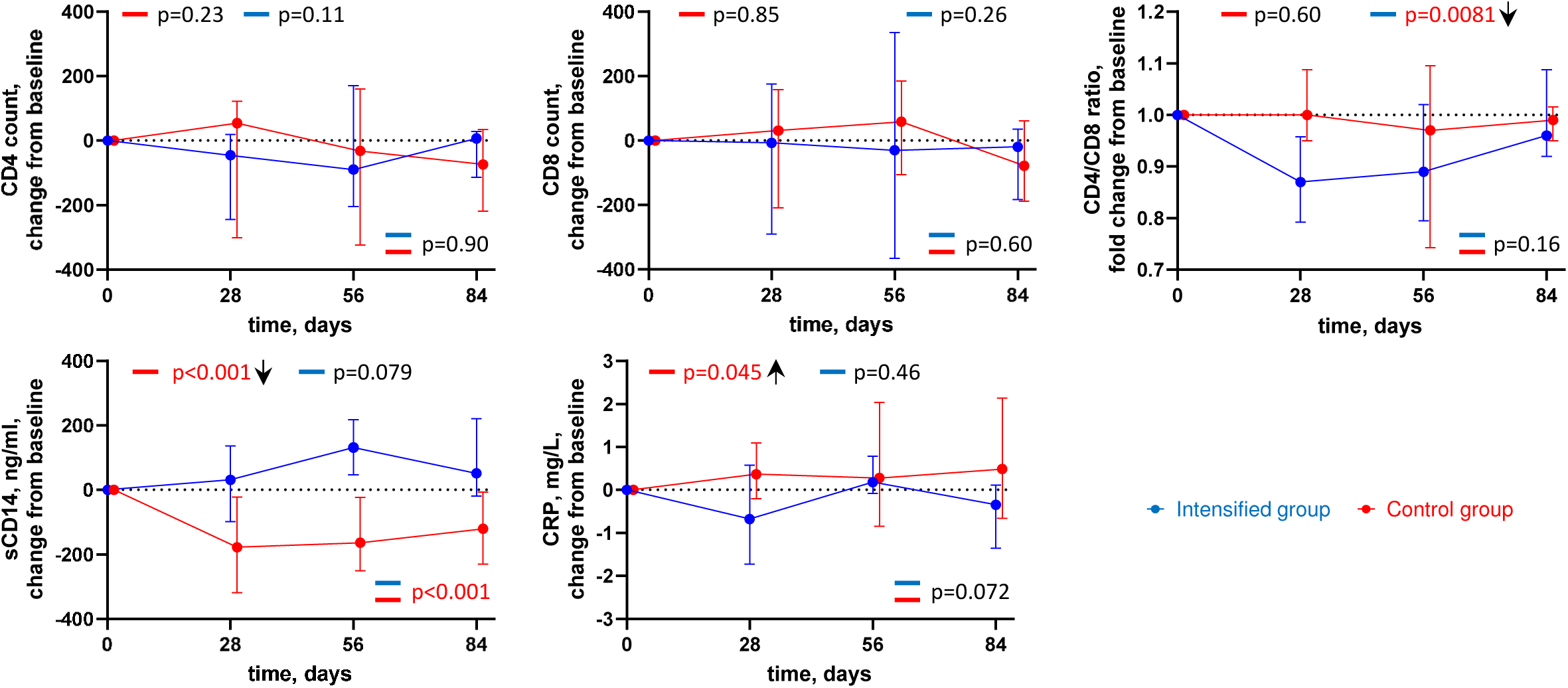
Longitudinal dynamics of clinical markers. Graphs show changes of markers from baseline on days 1, 28, 56, and 84 of the study in the intensified (blue) and control (red) groups. Median values and IQRs are shown. Linear mixed-effects modelling was used to calculate statistical significance. P values at the bottom of the graphs show the significance of between-group comparisons and those on top of the graphs show the significance of comparisons of the changes from baseline with zero in each group separately (intercept-only analysis). An upward or downward facing arrow next to a p value indicates a statistically significant increase or decrease from baseline, respectively. P values are marked red if significant (<0.05).

### Correlations between the measured parameters

Finally, we assessed pairwise correlations between the measured parameters at baseline, as well as between their time-weighted changes from baseline during the study (time-weighted areas under curve) and between their relative changes between days 0 and 84 of the study (Figures S9-S11). At baseline, we found strong positive pairwise correlations between the levels of different plasma inflammatory cytokines, as well as between the percentages of peripheral CD8+ T cells expressing TIGIT and inflammatory cytokine levels in plasma and tissue. Of the latter, especially strong correlations were observed between CD8+TIGIT+ cell percentages and plasma or tissue levels of IL-6 (*rho*=0.74 and *rho*=0.71, respectively; p<0.001). We also found positive correlations at baseline between US RNA/total DNA ratio and percentages of CD4+ T cells expressing CD38 and co-expressing CD38 and HLA-DR, markers of immune activation (*rho*=0.53, p=0.020 and *rho*=0.50, p=0.023, respectively). Furthermore, US RNA changes between days 0 and 84 positively correlated with changes in levels of expression of immune activation/exhaustion markers CD38+ on CD4+ cells and PD-1+ on CD8+ cells (*rho*=0.53, p=0.030 and *rho*=0.61, p=0.011, respectively) and of tissue inflammatory cytokine IL-1β mRNA (*rho*=0.62, p=0.0096). These correlations were even stronger for the changes between days 0 and 84 of US RNA/total DNA ratio (CD4+CD38+: *rho*=0.55, p=0.029; CD8+PD-1+: *rho*=0.73, p=0.0018; IL-1β mRNA: *rho*=0.76, p=0.0011). This confirms the results of our earlier study, where US RNA/total DNA ratio was found to correlate with markers of immune exhaustion (50). Interestingly, intact, but not total, HIV DNA in PBMCs strongly negatively correlated with time of viral suppression (*rho*=-0.67, p=0.0016), confirming earlier observations that intact HIV DNA decays on ART much faster than the defective (or total) HIV DNA (51, 52).

## Discussion

The aim of this study was to investigate whether ongoing viral replication contributes to HIV persistence under ART. To the best of our knowledge, this is the first study where ART intensification was achieved not by adding a new antiretroviral drug to the existing regimen but by increasing the dosage of a drug that was already part of the regimen pre-intensification. We evaluated the impact of doubling the dosage of DTG on HIV cellular and tissue reservoirs, immune activation, exhaustion, and inflammation in ART-suppressed PWH.

As expected, plasma and tissue DTG concentrations significantly increased between day 0 and day 84 in the intensified group but not in the control group. Accordingly, significant decreases in total HIV DNA, intact HIV DNA, US HIV RNA, and US RNA/total DNA ratio in PBMCs were observed during the study period in the intensified group but not in the control group. This strongly suggests that the pre-intensification ART regimen was not completely suppressive. The transient decrease in total DNA at day 28 may have been due to an initial decrease in newly infected cells upon ART intensification, however at the subsequent time points this effect was likely masked by proliferation (clonal expansion) of infected cells, mostly containing defective proviruses. In line with this, the number of intact proviruses decreased, but that of total proviruses did not change, between days 0 and 84 (Figures 2A, 2D).

In addition, intensification modestly yet significantly reduced percentages of CD4+ T cells expressing TIGIT, a T-cell exhaustion marker (53), and percentages of CD8+ T cells co-expressing CD38 and HLA-DR, T-cell activation markers. Interestingly, CD4+ cells expressing TIGIT have been shown to be enriched for HIV persistence in ART-treated PWH (54). Arguably, the reduction of HIV reservoir upon intensification has caused a corresponding reduction in chronic immune activation and exhaustion in this cohort. Indeed, changes in US HIV RNA levels and US RNA/total DNA ratio during the study positively correlated with changes in levels of markers immune activation, exhaustion, and inflammation, suggesting a link between changes in HIV reservoir activity and levels of immune activation and inflammation. These results are in line with previous studies reporting decreases in immune activation markers upon ART intensification (32–35).

In contrast, in the control group we measured significant longitudinal increases in total HIV DNA, percentages of CD4+ T cells expressing PD-1, another T-cell exhaustion marker, and in the plasma levels of IL-6, an inflammatory cytokine, and of CRP, another marker of inflammation. Residual HIV replication, among other factors, could have contributed to these effects in the control group. However, in the control group we also observed a reduction in the percentages of CD8+ T cells co-expressing a T-cell activation marker CD38 and a strong reduction in the plasma levels of sCD14. One possible explanation for these effects can be increased adherence of the participants to their ART regimens during the study.

Two previous studies have measured a transient increase in HIV episomal DNA (2-LTR circles) upon ART intensification with raltegravir, an INSTI (31, 36). As 2-LTR circles are a byproduct of HIV integration and are expected to accumulate if integration is blocked, this was interpreted as a strong evidence of new infections pre-intensification. However, another study did not observe any change in 2-LTR circles upon ART intensification with DTG (45). In our study, despite repeated attempts using different assays, we could only detect 2-LTR circles in a small minority of participants (<5%) (data not shown). Notably, the effects of intensification on 2-LTR circles in these previous studies were much more pronounced in participants who had been treated with a PI-based ART regimen pre-intensification (31, 36). As PI-based regimens have been shown to be less suppressive than other ART regimens (16, 55–57), they might have allowed for a higher level of new infections and consequently of 2-LTR circles in these studies.

In contrast, in our study, participants had been treated with a DTG-based regimen for at least two years and had had a suppressed plasma viral load (<50 copies/mL) for at least 2 years 11 months pre-intensification. Plausibly, the lack of detectable 2-LTR circles in our participants reflects the high potency of their ART regimens and a long period of virological suppression before the study. Remarkably, the intensification revealed that even this highly potent regimen was not completely suppressive. Thus, our results emphasize that 2-LTR circles are not a very sensitive marker of new infections due to their relative scarcity in infected cells. 2-LTR circles represent only ∼10% of total episomal DNA (58) and are not expected to be present in every newly infected cell. In contrast, a newly infected CD4+ T lymphocyte can contain hundreds to thousands of US HIV RNA copies on the peak of infection (59). Therefore, a change in the US RNA level upon ART intensification is a much more sensitive indicator of new infections than that of 2-LTR circles, as confirmed by this study.

Due to low penetration of antiretroviral drugs into tissue sites and anatomic sanctuaries (26–28, 60, 61), ART pressure on HIV replication is generally expected to be more relaxed in tissues than in peripheral blood. However, we did not observe any effect of the intensification on total HIV DNA in the rectal tissue. Although DTG concentration in the rectal tissue did increase upon intensification, it remained much lower than in the peripheral blood. This possibly contributed to the lack of detectable effect of intensification on the HIV DNA in tissue. Alternatively, intensification with DTG, an INSTI, had no effect in the tissue due to the preponderance of abortive infection events in lymphoid tissues that occur by cell-to-cell contact and are terminated prior to integration (62, 63). It is also possible that, similarly to the peripheral blood, the effect of the intensification on the reservoir in tissue was masked by the clonal expansion of defective proviruses by day 84 of the study.

The majority of previous studies that used INSTIs (raltegravir or DTG) as intensification drugs added it as a fourth drug using the standard dosage. For example, the DTG dosage in the study of Rasmussen et al. was 50 mg daily (45). In our study, we used a twice-higher dosage (100 mg daily). This DTG dosage is commonly used in clinical practice. Indeed, in PWH with confirmed or clinically suspected resistance to the integrase inhibitor class, the recommended dosage of DTG is 50 mg twice daily (https://www.cbip.be/fr/chapters/12?frag=10868). Based on our results (and pending the confirmation by larger studies), an increased dosage of DTG might be warranted in the future as a part of a standard regimen even in PWH without resistance. However, this has to be approached with caution as intensification has caused a transient decrease in the CD4/CD8 ratio in our study.

The main strength of our study is the measurement of a large number of virological and immunological parameters that allowed us to observe a significant impact of intensification on both virus and host. Our study is the first to observe significant reductions in as many as four HIV markers following ART intensification. Moreover, we measured plasma and tissue concentrations of DTG, allowing us to directly demonstrate their increase upon intensification. Our results are in line with some earlier studies that reported a decrease in cell-associated HIV RNA or a transient increase in 2-LTR circles upon intensification (31, 32, 36). However, they are in contrast with other studies, in which treatment intensification did not affect HIV markers (39, 40, 42). As this is the first ART intensification study to increase the dosage of an existing antiretroviral drug instead of adding a new drug, the results of this study and those of previous intensification studies cannot be directly compared.

Some limitations can be highlighted. First, we included twenty PWH (ten participants per arm). Although this small sample size does somewhat limit the robustness of the conclusions, we would like to emphasize that in this cohort, we performed a very detailed analysis, longitudinally measuring 30 biomarkers at five time points during ART. Secondly, despite the randomization, the groups were still not perfectly balanced as levels of total HIV DNA in PBMCs were by chance higher at baseline in the intensified group. However, this is unlikely to have affected the conclusions of the study because (i) this study has a within-subject (repeated-measures) design, with the longitudinal change of a parameter within the same participant during the study being the main outcome, and (ii) this difference most probably reflects a higher number of infected cells, and not a higher level of residual replication, at baseline in the intensified group. No significant differences between the groups were found at baseline in the levels of the other four virological markers we measured (US RNA in PBMCs, US RNA/total DNA ratio, intact DNA in PBMCs, and total DNA in tissue), as well as between the antiretroviral drug concentrations in plasma or in tissue. The lack of blinding and placebo control, the predominantly male study population, and the absence of post-intervention follow-up represent additional limitations of our study. Moreover, these findings should be considered exploratory and hypothesis-generating, warranting confirmation in larger, placebo-controlled, and blinded trials. Nevertheless, we believe that the convergence of the effect of intensification on multiple reservoir markers in the same direction indicates a potentially meaningful biological signal that merits further investigation.

In conclusion, doubling the DTG dosage in PWH who had been suppressed on DTG-containing ART for a number of years resulted in a reduction in the levels of four HIV reservoir markers in peripheral blood, as well as in markers of T-cell activation and exhaustion. If confirmed in larger clinical trials, these results could have an impact on the clinical management of PWH. Moreover, if ongoing viral replication does indeed replenish HIV reservoirs over time, then improving ART regimens should be a necessary part of the curative strategies.

## Methods

### Study design and participants

This was a phase 2 open-label, interventional, monocentric, randomized, and controlled clinical trial performed at the Hospital University of Liège (Belgium) in which 20 HIV-infected adults were enrolled. Eligible participants had been treated with a combination of 50 mg DTG, 600 mg ABC, and 300 mg 3TC for more than two years and the purpose of this study was to assess the impact of DTG intensification (an additional daily dose of 50 mg) compared to a control group that continued with the above regimen. This study was a proof-of-concept trial designed to reveal biological effects of ART intensification and the primary outcome was defined as “to evaluate the impact of treatment intensification at the level of total and replication-competent reservoir in blood and in tissues”, with a time frame of 3 months. Participants were required to have a plasma viral load suppressed below 20 HIV RNA copies/mL during the 12 months before screening (allowing “blips”) and an absolute CD4+ T lymphocyte count above 200 cells/mm^3^. One participant out of 20 had an isolated “blip” of 115 copies/mL nine months before the study initiation. The detailed inclusion and exclusion criteria can be found in the supplementary data (Supplementary data 1). The study was approved by the Liège University Hospital-Faculty Ethics Committee (Comité d’Éthique Hospitalo-Facultaire Universitaire de Liège, 2018/228). Prior to inclusion in the study, each participant had dated and signed an informed consent form. ClinicalTrials.gov identifier: NCT05351684.

Historical plasma HIV RNA measurements, CD4+ T-cell counts, and treatment data were retrieved from the outpatient medical records. The duration of continuous virological suppression was calculated as the duration of the latest period with undetectable plasma HIV RNA prior to the study initiation, allowing isolated ‘blips’ of 50–999 copies/ml. The duration of cumulative suppression was calculated by adding together all such periods of continuous suppression.

### Clinical procedures

Each participant was randomly assigned to the intensified group or to the control group. This study lasted 84 days in which 5 visits were programmed (days 0, 1, 28, 56, and 84). One visit prior to inclusion was added to perform a blood test to ensure that the participant met the inclusion/exclusion criteria for the study. The blood test included blood and platelet count, electrolyte panel, creatinine, LDH counts, liver tests, coagulation tests, CD4 and CD8 lymphocyte counts and a plasma viral load. β-HCG levels were measured in women at each visit to ensure that they were not pregnant. All participants completed the 12 week-long study and no individuals were lost to follow-up.

Safety and tolerability evaluations were conducted on day 28 and day 84 and included a clinical examination and routine laboratory testing (liver function tests, kidney function, and complete blood count). Medication adherence was also monitored through pill counts performed by the study nurses.

No virological blips above 50 copies/mL were observed and no adverse events were reported by participants during the 3-month intensification period. Although CPK levels were not included in the routine biological monitoring, no participant reported muscle pain or other symptoms suggestive of muscle toxicity.

Blood samples were collected in 4 EDTA tubes (∼40mL) at days 0, 1, 28, 56, and 84 and directly sent to the AIDS reference laboratory of Liège University Hospital for PBMC isolation. PBMC samples were stored at –150°C for measurements of the HIV reservoir and immunological parameters. Plasma samples were used to measure soluble inflammation markers. Ten flash-frozen rectal biopsies were collected on days 0 and 84 during a routine gastroenterology visit. The cryotubes containing the biopsies were directly placed in liquid nitrogen and stored at – 150°C.

### PBMC isolation

PBMCs were isolated from whole blood by density gradient centrifugation using Ficoll-Paque® Plus. The blood samples were collected in EDTA tubes and centrifuged at 1200 X g for 10 min (acceleration 9, brake 9). The plasma was removed and stored at –150°C. The blood was then well homogenized with RPMI 1640, gently transferred in a Falcon tube that contained 12,5 mL of Ficoll, and centrifuged at 700 X g for 20 minutes (acceleration 6, brake 0). The PBMC layer was removed, diluted in 50 mL RPMI 1640, and centrifuged at 300 X g for 7 minutes (acceleration 9, brake 9). The supernatant was removed, and the process was repeated two more times, completing three washes. The pellet was resuspended, and the cells were counted. Foetal bovine serum (FBS) was added to the re-suspended pellet to obtain 10×10^6^ cells/500 μL. For cryopreservation, 500 μL of freeze mixture (10% DMSO in FBS) was added to 500 μL of the cell suspension in the cryotubes. Cryotubes were slowly frozen in the Mr. Frosty™ freezing container at –80 °C and transferred to –150°C the next day.

### Quantification of total HIV DNA and cell-associated US HIV RNA in PBMCs

Total nucleic acids were extracted from 2×10^6^ PBMCs using Boom isolation method (64). Total HIV DNA was measured by qPCR using the previously described primer/probe combination that amplifies the HIV packaging signal (Ψ) region (49). Extracted cellular RNA was treated with DNase (DNA-free kit; ThermoFisher Scientific) to remove genomic DNA that could interfere with the quantitation and reverse transcribed into cDNA using random primers and SuperScript III reverse transcriptase (all from ThermoFisher Scientific). To quantify cell-associated US HIV RNA, this cDNA was pre-amplified using primer pair Ψ_F (49) and HIV-FOR (65). The product of this PCR was used as template for a seminested qPCR with the Ψ primer/probe combination (49). HIV DNA or RNA copy numbers were determined using a 7-point standard curve with a linear range of more than 5 orders of magnitude that was included in every qPCR run and normalized to the total cellular DNA (by measurement of β-actin DNA) or RNA (by measurement of 18S ribosomal RNA) inputs, respectively, as described previously (66). Non-template control wells were included in every qPCR run and were consistently negative. Total HIV DNA and US RNA were detectable in 90.7% and 66.7% of the samples, respectively. Undetectable measurements of US RNA or total DNA were assigned the values corresponding to 50% of the corresponding assay detection limits, with a maximum of 25 copies/μg total RNA or 50 copies/10^6^ PBMCs, respectively. The detection limits depended on the amounts of the normalizer (input cellular DNA or RNA) and therefore differed among samples. Measurements with low input cellular DNA or RNA and undetectable total HIV DNA or US RNA (n=2 and n=3, respectively), were excluded from the analysis. HIV transcription levels per provirus (US RNA/total DNA ratios) were calculated taking into account that 10^6^ PBMCs contain 1 μg of total RNA (67).

### Quantification of intact HIV DNA in PBMCs

Intact HIV DNA was quantified on days 0 and 84 of the study by the IPDA (49). In brief, genomic DNA was isolated from 4×10^6^ PBMCs using Puregene Cell Kit (QIAGEN Benelux B.V.) according to the manufacturer’s instructions and digested with *Bgl*I restriction enzyme (ThermoFisher Scientific) as described previously (68). Notably, only a small minority (<8%) of HIV clade B sequences contain *Bgl*I recognition sites between Ψ and *env* amplicons, therefore *Bgl*I digestion is not expected to substantially influence the IPDA output, while improving the assay sensitivity by increasing the genomic DNA input into a ddPCR reaction (68). After desalting by ethanol precipitation, genomic DNA was subjected to two separate multiplex droplet digital PCR (ddPCR) assays: one targeting HIV Ψ and *env* regions using primers and probes described previously, including the unlabelled *env* competitor probe to exclude hypermutated sequences (49), and one targeting the cellular *RPP30* gene, which was measured to correct for DNA shearing and to normalize the intact HIV DNA to the cellular input. The *RPP30* assay amplified two regions, with amplicons located at exactly the same distance from each other as HIV Ψ and *env* amplicons. The first region was amplified using a forward primer 5’-AGATTTGGACCTGCGAGCG-3’, a reverse primer 5’-GAGCGGCTGTCTCCACAAGT-3’, and a fluorescent probe 5’-FAM-TTCTGACCTGAAGGCTCTGCGCG-BHQ1-3’ (69). The second region was amplified using a forward primer 5’-AGAGAGCAACTTCTTCAAGGG-3’, a reverse primer 5’-TCATCTACAAAGTCAGAACATCAGA-3’, and a fluorescent probe 5’-HEX-CCCGGCTCTATGATGTTGTTGCAGT-BHQ1-3’. The ddPCR conditions were as described previously (49) with some minor amendments: we used 46 cycles of denaturation/annealing/extension and the annealing/extension temperature was 60°C. Intact HIV DNA was detectable in 79.5% of the samples. Undetectable measurements of intact HIV DNA were assigned the values corresponding to 50% of the assay detection limits. The detection limits depended on the amounts of the normalizer (input cellular DNA) and therefore differed among samples. For two participants, in whom intact HIV DNA on day 0 was undetectable due to a technical issue, we included day 1 values in the analysis. Excluding these two participants did not change the conclusions (data not shown).

### Quantification of total HIV DNA in rectal biopsies

Prior to total HIV DNA isolation, rectal biopsies were homogenized with the TissueLyser (QIAGEN Benelux B.V.). Total HIV DNA was isolated using ReliaPrep™ gDNA Tissue Miniprep System (Promega) according to the manufacturer’s instructions. Total HIV DNA in rectal biopsies was quantified using exactly the same method as total HIV DNA in PBMCs (see above). Total HIV DNA was detectable in 92.5% of the rectal tissue samples. Undetectable measurements of total HIV DNA were assigned the values corresponding to 50% of the corresponding assay detection limits. The detection limits depended on the amounts of the normalizer (input cellular DNA) and therefore differed among samples.

### Quantification of immune activation and exhaustion

Flow cytometry was used to quantify surface markers of immune activation and exhaustion (CD38, HLA-DR, PD-1, TIGIT) on CD4+ and CD8+ T cells. Cryopreserved PBMCs (10×10^6^ cells) were thawed and stained with a fixable viability dye Zombie NIR (Sony), and the following antibodies: CD3-APC/Fire810, CD4-PE/Fire700, CD8a-PercP (all from Sony), PD-1-PE, TIGIT-BV421, HLA-DR-BB515, CD38-BV711 (all from BD Biosciences). Cells were then washed, fixed in 1% paraformaldehyde, and run on a FACS SONY ID7000. Analysis was performed using FlowJo software (v10.8.1). Cells were gated according to morphological parameters (forward and side scatter) and viability. The complete gating strategy can be found in the supplemental data (Fig. S12). To exclude the effect of inter-assay variation on the data, samples from each participant were always analysed together.

### Quantification of systemic and tissue inflammation

To quantify systemic inflammation, the levels of five plasma inflammatory cytokines (IFN-γ, IL-1β, IL-6, IL-17α, TNF-α) were measured on MAGPIX (Luminex) using the ProcartaPlex™ Human Inflammation Panel, 20plex (ThermoFisher Scientific). sCD14 was quantified by Human CD14 Quantikine ELISA Kit (R&D Systems). CRP was measured by high-sensitivity C-reactive protein (hs-CRP) test.

To measure tissue inflammation, we determined rectal tissue levels of mRNAs encoding five inflammatory cytokines (IL-1β, IL-6, IFN-γ, IL-17 α, and TNF-α). Rectal tissue biopsies were manually homogenized and lysed in buffer L6 (64) and RNA was extracted using Boom isolation method (64). Extracted RNA was treated with DNase (DNA-free kit; ThermoFisher Scientific) and reverse transcribed using random primers and SuperScript III reverse transcriptase (all from ThermoFisher Scientific). Cytokine mRNAs were quantified using TaqMan Gene Expression Assays (all from ThermoFisher Scientific): IL-1β – Hs01555410_m1, IL-6 – Hs00174131_m1, IFN-γ – Hs00989291_m1, IL-17 α – Hs00174383_m1, TNF-α – Hs00174128_m1. Levels of cytokine mRNAs were normalized to that of *GAPDH* mRNA, measured using a TaqMan Gene Expression Assay Hs02758991_g1.

### Quantification of antiretroviral drug concentrations

Briefly, DTG was extracted from plasma samples via methanolic protein precipitation and from tissue samples via tissue homogenization, followed by protein precipitation. A DTG standard calibration curve over the range 20.0-5,000 ng/mL and stable isotope labelled internal standard were used to generate an analyte/internal standard response. Analyte/internal standard response was detected by liquid chromatography tandem mass spectrometry (LCMS) based on previously described methods (26, 60, 70). Tissue penetration ratios of DTG were calculated as the ratios of DTG concentrations in tissue (as ng/mL after conversion from ng/g by dividing the values in ng/g by 1.06 (71)) to the concentrations in plasma (ng/mL). Lamivudine (3TC) concentrations were determined with an LCMS method similar to our previously described procedures (72). Differences are as follows: 3TC stock and standard working solutions were prepared in methanol and demonstrated similar stability. The MS used was a triple-quadrupole AB Sciex 5500 series monitoring positive ion transitions 230.1 –> 112.1 (3TC) and 233.1 –> 115.1 (stable labeled internal standard). Mobile phase was 12.5% acetonitrile and 0.1% formic acid in deionized water. No source rinsing mobile phase was needed. The limit of quantitation was 30 ng/mL and the upper bound was 6,000 ng/mL. Total accuracy ranged from 94.3–96.9% for. Within-day coefficients of variation were below 5%, between-day coefficients of variation were below 5%. Stability of 3TC in human plasma was also similar.

### Statistical analysis

Baseline parameters were compared using Mann-Whitney tests for continuous variables and Fisher’s exact tests for categorical variables. Virological parameters were log_10_-transformed before analysis. Longitudinal dynamics of the measured parameters was modelled using linear mixed-effects analysis. This analysis was used both to compare the longitudinal changes from baseline between the intensified and the control groups and to compare the changes from baseline with zero in each of these groups separately (intercept-only analysis). As a sensitivity analysis, we performed a comparison between models that do and do not include time as a covariate. Including time in the models did not change the conclusions (Supplementary Tables 1 and 2). Paired Wilcoxon tests were used for within-group comparisons of parameters between days 0 and 84. For these comparisons, pairs where both values were undetectable were excluded from the analysis. Time-weighted changes from baseline were calculated by dividing the areas under curve (computed using the trapezoid rule) by the time period of the study. Time-weighted changes from baseline were not calculated if the baseline value and one or more of the subsequent values were undetectable. Time-weighted changes from baseline and ratios of the parameters between different study days were compared between the groups using Mann-Whitney tests. Spearman tests were used to produce correlation matrices of the parameters at baseline, as well as of time-weighted changes from baseline and of changes between day 0 and day 84 of the measured parameters. Changes of the parameters from baseline, as well as US RNA/total DNA ratios, were not calculated if both values in a pair were undetectable. Data were analysed using Prism 10.2.0 (GraphPad Software) or IBM SPSS Statistics 28.0.1.0. All statistical tests were two-sided, and p values of <0.05 were considered statistically significant.

## Data Availability

All data supporting the findings of this study are available within the paper and its Supplementary Information.

## Acknowledgements

We are grateful to Marine Hubert and Raquel Mateus for technical assistance in PBMC isolation. We acknowledge the participation and commitment of the trial participants who made the study possible. C.F.L. is a FRIA grantee of the Belgian Fund for Scientific Research (Fonds de la Recherche Scientifique –FNRS) and is supported by the Fondation Léon Fredericq. This work was supported in part by grant R01-AI124965 (to C.V.F.) from the US National Institutes of Health. A.O.P. acknowledges grant support from amfAR, The Foundation for AIDS Research (grant no. 1110680–77-RPRL), and from Partnership NWO-Dutch AIDS Fonds ‘HIV cure for everyone’ (grant no. KICH2.V4P.AF23.001). G.D. is supported by the Belgian Fund for Scientific Research (Fonds de la Recherche Scientifique –FNRS).

